# Vascular responses to drug-coated balloon angioplasty in lipid-rich vs non-lipid-rich coronary lesions

**DOI:** 10.1101/2025.07.07.25331064

**Authors:** Yutaka Matsuhiro, Yasuyuki Egami, Hiroaki Nohara, Shodai Kawanami, Koji Yasumoto, Naotaka Okamoto, Yasuharu Matsunaga-Lee, Masamichi Yano, Daisuke Nakamura, Isamu Mizote, Yasushi Sakata, Masami Nishino

**Affiliations:** Department of Cardiovascular Medicine, Osaka University Graduate School of Medicine, Osaka, Japan; Division of Cardiology, Osaka Rosai Hospital, Osaka, Japan

**Author notes:** **Corresponding Author:** Masami Nishino, MD, PhD, FACC, FESC Division of Cardiology, Osaka Rosai Hospital, 1179-3, Nagasone-cho, Sakai-city, Osaka 591-8025, Japan.

**Keywords:** lipid-rich plaque, drug-coated balloon, NIRS-IVUS

## Abstract

**BACKGROUND:** Lipid-rich plaques are strongly associated with adverse cardiac events. However, the vascular response of lipid-rich lesions to drug-coated balloon (DCB) angioplasty has not been fully elucidated. This study aimed to compare vascular responses following DCB angioplasty between lipid-rich and non-lipid-rich coronary lesions.

**METHODS:** This study included patients scheduled for DCB angioplasty for de novo small vessel lesions between February 2021 and April 2024. All patients underwent pre- and post-procedural near-infrared spectroscopy combined with intravascular ultrasound (NIRS-IVUS), along with follow-up coronary angiography and NIRS-IVUS at 8 months. Lesions were categorized as lipid-rich or non-lipid-rich based on the pre-procedural maximum lipid core burden index over 4mm (maxLCBI_4_mm). Changes in plaque volume and maxLCBI_4_mm from post-procedure to the 8-month follow-up, as well as the incidence of late lumen enlargement (LLE), were compared between the two groups.

**RESULTS:** Forty-six patients were included in the final analysis: 15 in the lipid-rich group and 31 in the non-lipid-rich group. LLE occurred in 27% of the lipid-rich group and 39% of the non-lipid-rich group (P = 0.52). Percent plaque volume showed minimal change in the lipid-rich group (+0.3%), while a significant reduction was observed in the non-lipid-rich group (−3.6%; P = 0.045). Conversely, maxLCBI_4_mm significantly decreased in the lipid-rich group compared with the non-lipid-rich group (−240 vs. −42; P = 0.0030), resulting in comparable values at the 8-month follow-up.

**CONCLUSIONS:** Despite limited plaque regression, lipid-rich lesions demonstrated favorable vascular responses, including LLE and plaque stabilization, following DCB angioplasty.

This study was registered in the UMIN Clinical Trials Registry (UMIN000043158). https://www.umin.ac.jp/

**What is Known:**

- Vascular responses, such as vessel enlargement and plaque regression, are frequently observed following drug-coated balloon (DCB) angioplasty, contributing to late lumen enlargement (LLE).

**What the Study Adds:**

- Plaque regression is less likely to occur in lipid-rich lesions.
- LLE occurs at a similar rate between lipid-rich and non-lipid-rich lesions.
- Lipid-rich lesions show a significant reduction in lipid burden.

## Introduction

Recent large-scale trials have demonstrated that drug-coated balloon (DCB) angioplasty for de novo coronary artery disease provides clinical outcomes comparable to those of drug-eluting stents, particularly in small vessel lesions.^1,2^ Moreover, DCB angioplasty offers several specific advantages, including the avoidance of stent thrombosis associated with stent malapposition or neoatherosclerosis following stent implantation.^3^ In addition, favorable vascular responses, such as vessel enlargement and plaque regression, are frequently observed following DCB angioplasty, contributing to late lumen enlargement (LLE).^4,5^ However, the relationship between baseline lesion characteristics and both vascular responses and changes in plaque composition following DCB angioplasty remains unclear. Near-infrared spectroscopy combined with intravascular ultrasound (NIRS-IVUS) is a promising imaging modality for identifying high-risk, lipid-rich plaques through simple and objective quantification using maximum lipid core burden index over 4mm (maxLCBI_4_mm).^6^ Lipid-rich plaques, widely recognized as vulnerable plaques, are strongly associated with adverse cardiac events.^7^ The management of lesions containing such plaques is currently a topic of active discussion.^8^ Therefore, the objective of this study was to compare vascular responses and changes in plaque composition following DCB angioplasty between lipid-rich and non-lipid-rich lesions, as assessed by NIRS-IVUS.

## Methods

### STUDY DESIGN AND POPULATION

The SWITCH (**<u>S</u>**tent-less Percutaneous Coronary Intervention **<u>w</u>**ith **<u>I</u>**maging **<u>T</u>**e**<u>ch</u>**nology) study was a single-center, prospective interventional study. The study enrolled consecutive patients scheduled for DCB angioplasty for de novo small vessel coronary artery disease at Osaka Rosai Hospital between February 2021 and April 2024. All patients underwent pre- and post-procedural NIRS-IVUS, along with follow-up coronary angiography and NIRS-IVUS at 8 months. In Japan, at the time of the study’s initiation, only small-sized DCBs (less than 3mm in diameter) are approved for de novo lesions. ^9^ Therefore, large-vessel lesions were excluded from this study. All procedures in this study were performed using the SeQuent Please NEO (B. Braun, Melsungen, Germany). The exclusion criteria were as follows: 1) large-vessel lesions requiring large DCBs (≥ 3 mm); 2) acute coronary syndrome; 3) patients undergoing hemodialysis; 4) ostial lesions or in-stent restenosis; 5) patients who required stent implantation due to unsuccessful lesion preparation prior to DCB angioplasty; 6) patients who underwent target lesion revascularization during the 8-month periods; and 7) patients who did not undergo follow-up NIRS-IVUS. This study was conducted in accordance with the principles of the Declaration of Helsinki and was approved by the Institutional Review Board of Osaka Rosai Hospital. Written informed consent was obtained from all patients prior to their participation in the procedure and study enrollment. All data analyses, including angiographic and IVUS assessments, were performed at Osaka University; therefore, the study was also approved by the Institutional Review Board of Osaka University. This study was registered in the UMIN Clinical Trials Registry (UMIN000043158).

### PROCEDURES

All procedures, including pre dilatation and the use of atherectomy devices, were underwent under IVUS guidance, and were left to the discretion of the individual operators. The NIRS-IVUS system with a 3.2 Fr rapid exchange catheter, auto-pullback controller, and console (Makoto™ Intravascular Imaging System, Infraredx, Inc., Bedford, MA, USA) was used in this study. The DCB strategy was implemented in accordance with the recommendations of the International DCB Consensus Group.^10^ The use of scoring or cutting balloons with a ballon-to-artery ratio of 1.0, as well as DCBs of the same size as the pre-dilatation balloons, was also recommended in this study.^9,10^ Bail-out stent implantation was permitted in the following situations after lesion preparation; 1) thrombolysis in myocardial infarction flow grade < 3; 2) coronary dissection classified as the National Heart, Lung and Blood Institute classification ≥ Type D on coronary angiography; or 3) residual diameter stenosis > 30% on coronary angiography or area stenosis ≥ 50% on IVUS.^9,10,11,12^

### IMAGING ANALYSIS

All obtained angiographic and NIRS-IVUS data were transferred to the imaging core laboratory at Osaka University for analysis. The core laboratory analyzed the angiograms using QAngio XA Software (version 8.0, Medis Medical Imaging Systems, Leiden, the Netherlands) and the NIRS-IVUS data using QIvus Software (version 3.1, Medis Medical Imaging Systems, Leiden, the Netherlands). Severe calcification on pre-procedural IVUS and medial dissection > 90 degrees on post-procedural IVUS were assessed as part of the qualitative analysis.^13,14^ For quantitative analysis, IVUS images were evaluated at 1-mm intervals throughout the DCB-treated segment, both immediately after DCB angioplasty and at the 8-month follow-up. Vessel volume, lumen volume, plaque volume and percent plaque volume were measured in accordance with standard IVUS analysis protocols.^15^ maxLCBI_4_mm was obtained from pre-procedural, post-procedural and follow-up IVUS. The lipid plaque volume was calculated as the portion of the total plaque volume corresponding to areas identified as lipid-containing by NIRS-IVUS. The non-lipid plaque volume was calculated by subtracting the lipid plaque volume from the total plaque volume. To account for lesion length, all volume parameters were adjusted and expressed as mean volume per unit length. Changes in all parameters were calculated as the values at the 8-month follow-up minus those immediately after the procedure.

### STUDY ENDPOINTS AND DEFINITIONS

Patients were divided into two groups, lipid-rich and non-lipid-rich, based on pre-procedural maxLCBI_4_mm, using a cutoff value of > 400, as established in the LRP study.^7^ From the post-procedure to the 8-month follow-up, serial changes in each volumetric parameter and maxLCBI_4_mm, as well as the incidence of LLE, were compared between the two groups. LLE was defined as the ratio of the mean lumen area at follow-up to that immediately after the procedure, with a value ≥ 1.1, as defined in a previous study.^14^

### STATISTICAL ANALYSIS

Categorical variables were compared using the chi-square or Fisher’s exact test, as appropriate, and presented as numbers (percentages). Continuous variables were compared using a Student’s *t*-test and the Mann-Whitney *U*-test, based on the distribution and stated as the mean ± standard deviation or median [interquartile range; Q1, Q3]. All clinical, angiographic, and NIRS-IVUS data were compared between the lipid-rich and the non-lipid-rich groups. In addition, multivariate logistic regression analysis was performed to identify significant predictors of LLE. Covariates included relevant clinical and NIRS-IVUS variables that were significantly associated with the outcome of interest in the univariable analysis (p < 0.10). Statistical significance was defined as p <0.050. A sample size calculation was not conducted because of the lack of prior research to inform parameter estimates. Statistical analyses were performed using R software (version 4.3.2; R Foundation for Statistical Computing, Vienna, Austria; http://www.r-project.org/).

## Results

A total of 76 consecutive patients were scheduled to undergo DCB angioplasty with NIRS-IVUS during the study period. After one case of bail-out stenting was excluded, 75 patients successfully underwent DCB angioplasty. During the 8-month follow-up period, one case of target lesion revascularization occurred. One patient was excluded due to poor IVUS image quality. In addition, NIRS-IVUS was not performed in 27 patients for the following reasons: 20 patients due to restrictions during the COVID-19 pandemic, 2 based on physician judgment, 3 due to angiographically severe stenosis, and 2 due to poor prognosis from non-cardiac diseases. As a result, a total of 46 patients were included in the final analysis: 15 in the lipid-rich group and 31 in the non– lipid-rich group. (Figure 1)

**Figure 1.**
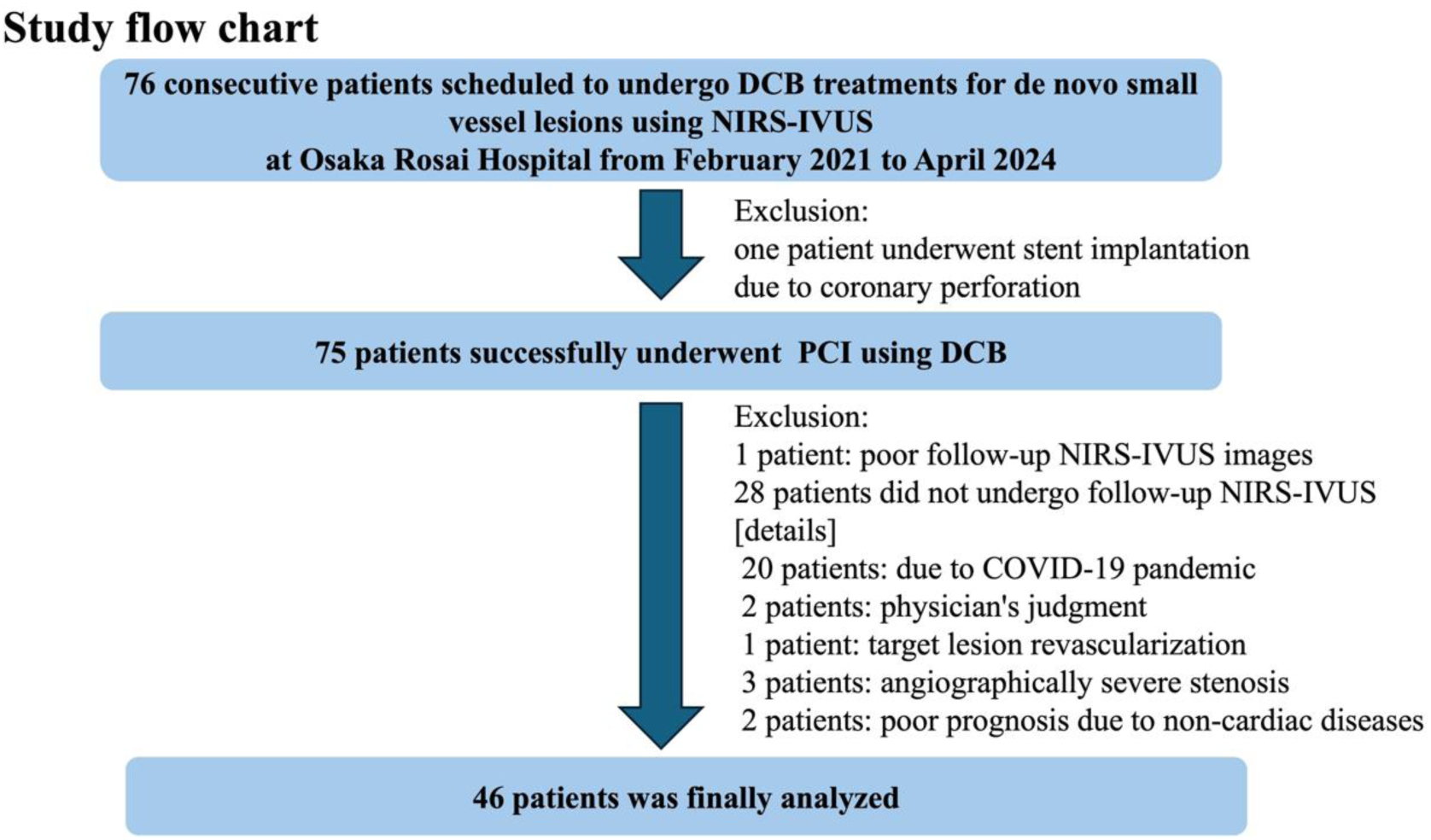
Study flow chart. Of the 76 patients scheduled for DCB angioplasty with NIRS-IVUS, 75 successfully underwent the procedure. After excluding one bail-out stenting and one target lesion revascularization case, 46 patients (15 lipid-rich, 31 non–lipid-rich) were included in the final analysis. DCB = drug-coated balloon; NIRS-IVUS = near-infrared spectroscopy combined with intravascular ultrasound.

### PATIENT CHARACTERISTICS

Baseline patient characteristics are shown in Table 1. The mean age was 74 years, and 87 % of the overall cohort were male. The prevalence of hypertension was higher in the lipid-rich group than in the non-lipid-rich group. Other comorbidities, including dyslipidemia and diabetes mellitus, were similarly observed between the two groups. A history of prior coronary intervention was common in both groups (80 % in the lipid-rich group vs. 71 % in the non-lipid-rich group; P = 0.72). The prescription rate of direct oral anticoagulants was higher in the lipid-rich group, possibly due to a higher prevalence of atrial fibrillation compared with the non-lipid-rich group. Regarding lipid-lowering therapy, statin use was high, and the median low-density lipoprotein cholesterol level was approximately 60 mg/dL in both groups at baseline and at the 8-month follow-up.

**Table 1.**
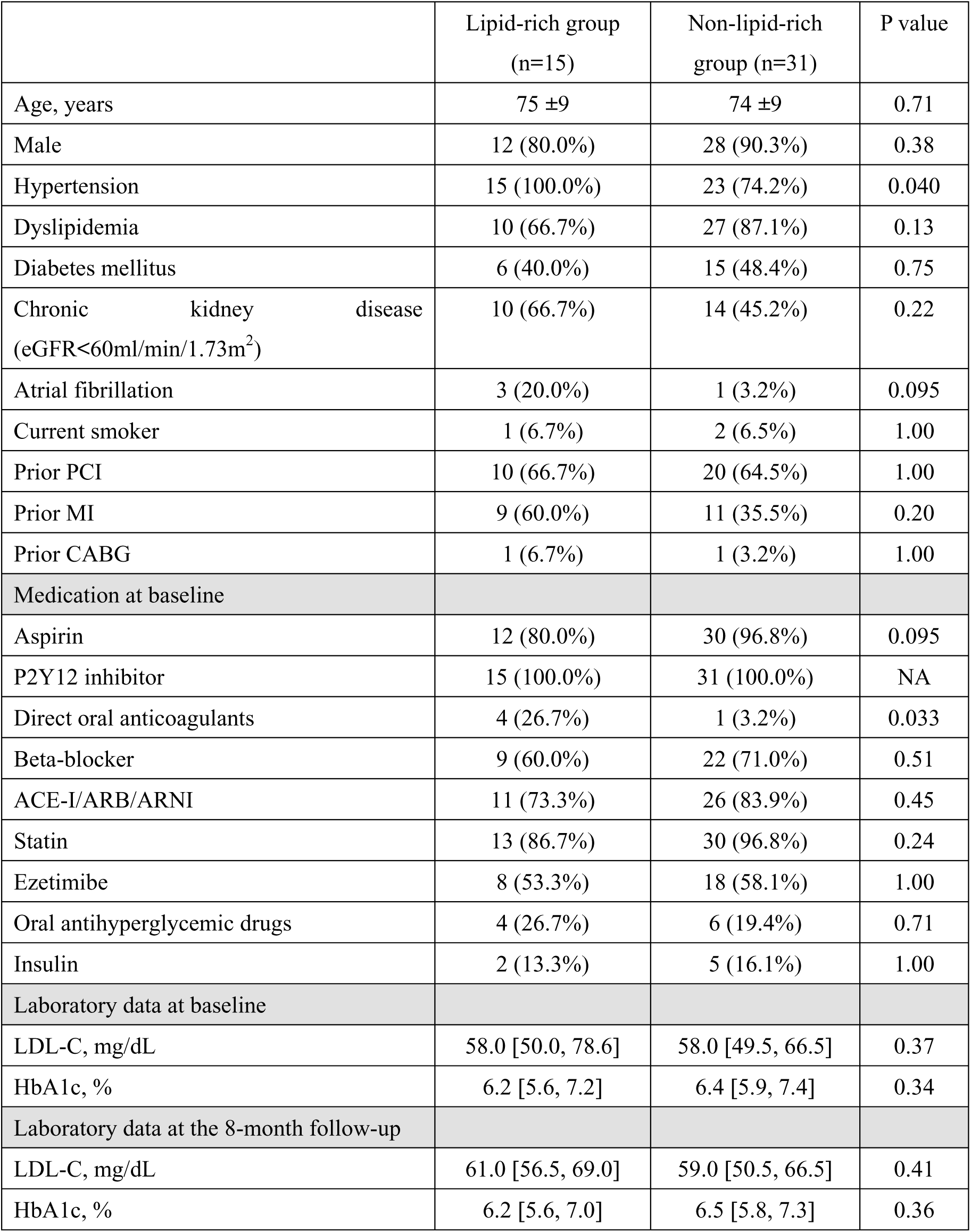
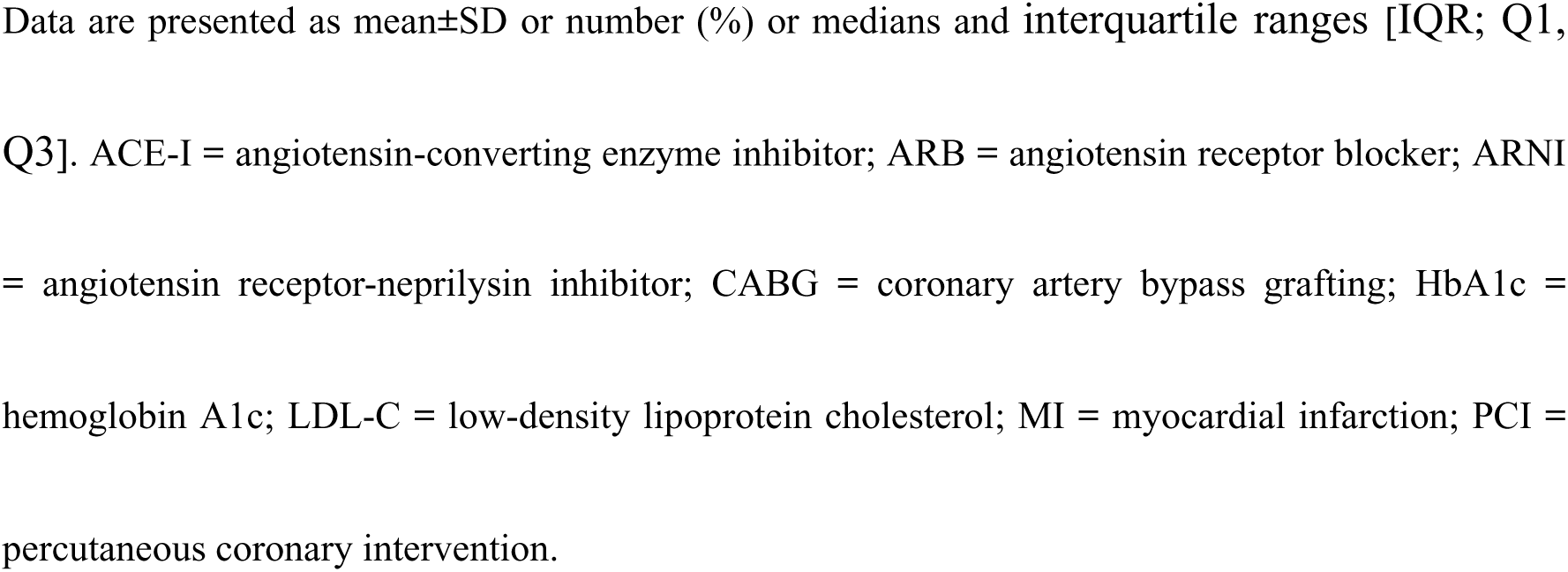
Baseline characteristics.

|  | Lipid-rich group<br>(n=15) | Non-lipid-rich<br>group (n=31) | P value |
| --- | --- | --- | --- |
| Age, years | 75 ±9 | 74 ±9 | 0.71 |
| Male | 12 (80.0%) | 28 (90.3%) | 0.38 |
| Hypertension | 15 (100.0%) | 23 (74.2%) | 0.040 |
| Dyslipidemia | 10 (66.7%) | 27 (87.1%) | 0.13 |
| Diabetes mellitus | 6 (40.0%) | 15 (48.4%) | 0.75 |
| Chronic kidney disease<br>(eGFR<60ml/min/1.73m<sup>2</sup>) | 10 (66.7%) | 14 (45.2%) | 0.22 |
| Atrial fibrillation | 3 (20.0%) | 1 (3.2%) | 0.095 |
| Current smoker | 1 (6.7%) | 2 (6.5%) | 1.00 |
| Prior PCI | 10 (66.7%) | 20 (64.5%) | 1.00 |
| Prior MI | 9 (60.0%) | 11 (35.5%) | 0.20 |
| Prior CABG | 1 (6.7%) | 1 (3.2%) | 1.00 |
| Medication at baseline |  |  |  |
| Aspirin | 12 (80.0%) | 30 (96.8%) | 0.095 |
| P2Y12 inhibitor | 15 (100.0%) | 31 (100.0%) | NA |
| Direct oral anticoagulants | 4 (26.7%) | 1 (3.2%) | 0.033 |
| Beta-blocker | 9 (60.0%) | 22 (71.0%) | 0.51 |
| ACE-I/ARB/ARNI | 11 (73.3%) | 26 (83.9%) | 0.45 |
| Statin | 13 (86.7%) | 30 (96.8%) | 0.24 |
| Ezetimibe | 8 (53.3%) | 18 (58.1%) | 1.00 |
| Oral antihyperglycemic drugs | 4 (26.7%) | 6 (19.4%) | 0.71 |
| Insulin | 2 (13.3%) | 5 (16.1%) | 1.00 |
| Laboratory data at baseline |  |  |  |
| LDL-C, mg/dL | 58.0 [50.0, 78.6] | 58.0 [49.5, 66.5] | 0.37 |
| HbA1c, % | 6.2 [5.6, 7.2] | 6.4 [5.9, 7.4] | 0.34 |
| Laboratory data at the 8-month follow-up |  |  |  |
| LDL-C, mg/dL | 61.0 [56.5, 69.0] | 59.0 [50.5, 66.5] | 0.41 |
| HbA1c, % | 6.2 [5.6, 7.0] | 6.5 [5.8, 7.3] | 0.36 |

Data are presented as mean ± SD or number (%) or medians and interquartile ranges [IQR; Q1, Q3]. ACE-I = angiotensin-converting enzyme inhibitor; ARB = angiotensin receptor blocker; ARNI = angiotensin receptor-neprilysin inhibitor; CABG = coronary artery bypass grafting; HbA1c = hemoglobin A1c; LDL-C = low-density lipoprotein cholesterol; MI = myocardial infarction; PCI = percutaneous coronary intervention.

### LESION, PROCEDURAL, AND ANGIOGRAPHIC CHARACTERISTICS

Lesion and procedure characteristics, as well as quantitative coronary angiography (QCA) data, are presented in Table 2. Scoring or cutting balloons were used in 93% of all procedures. The use of debulking devices tended to be higher in the lipid-rich group than in the non-lipid-rich group. Lesions in the lipid-rich group tended to be longer lesion than those in the non-lipid-rich group, resulting in the use of longer DCB. Percent diameter stenosis did not differ between the two groups at baseline, post-procedure, and at the 8-month follow-up in the QCA analysis.

**Table 2.**
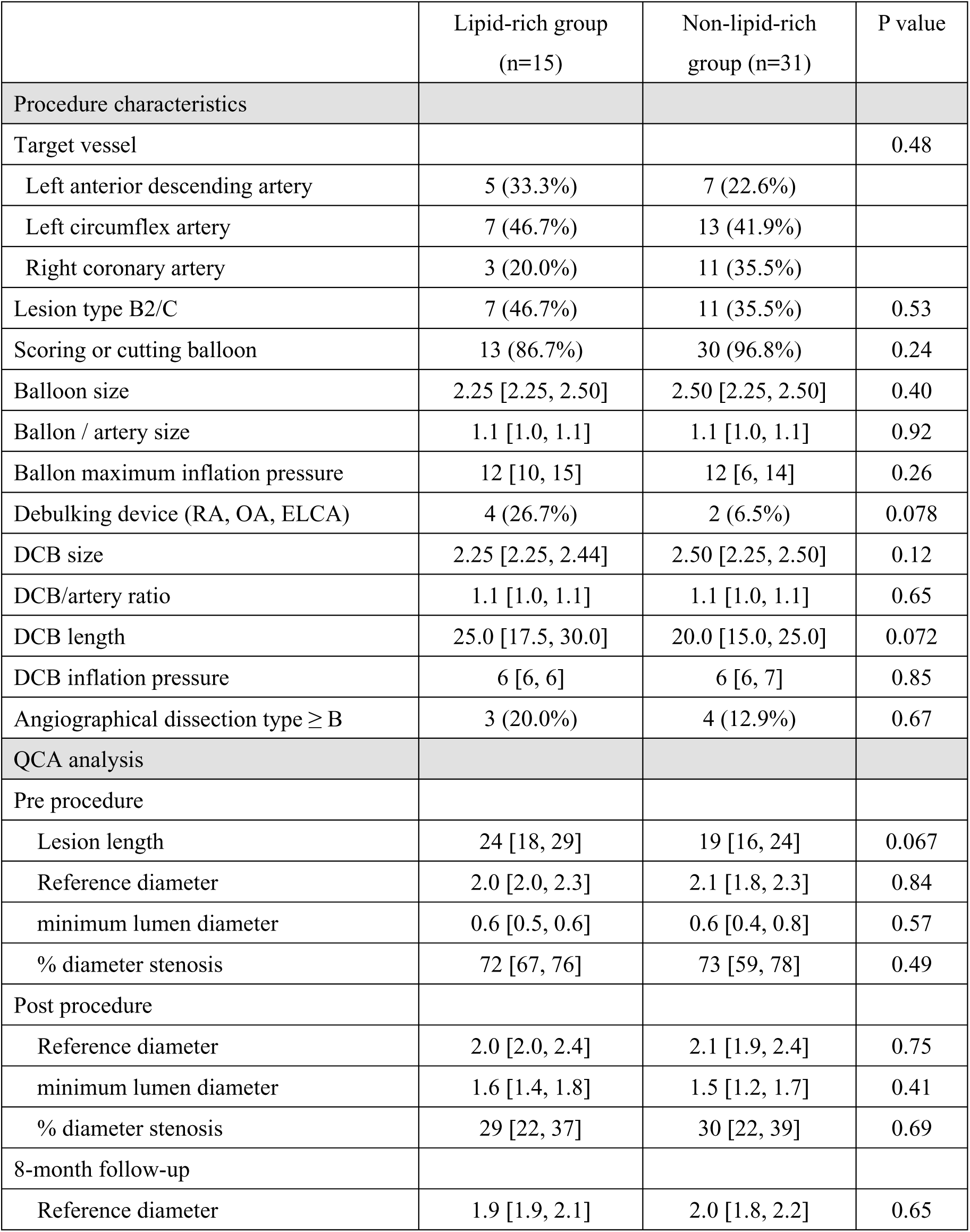
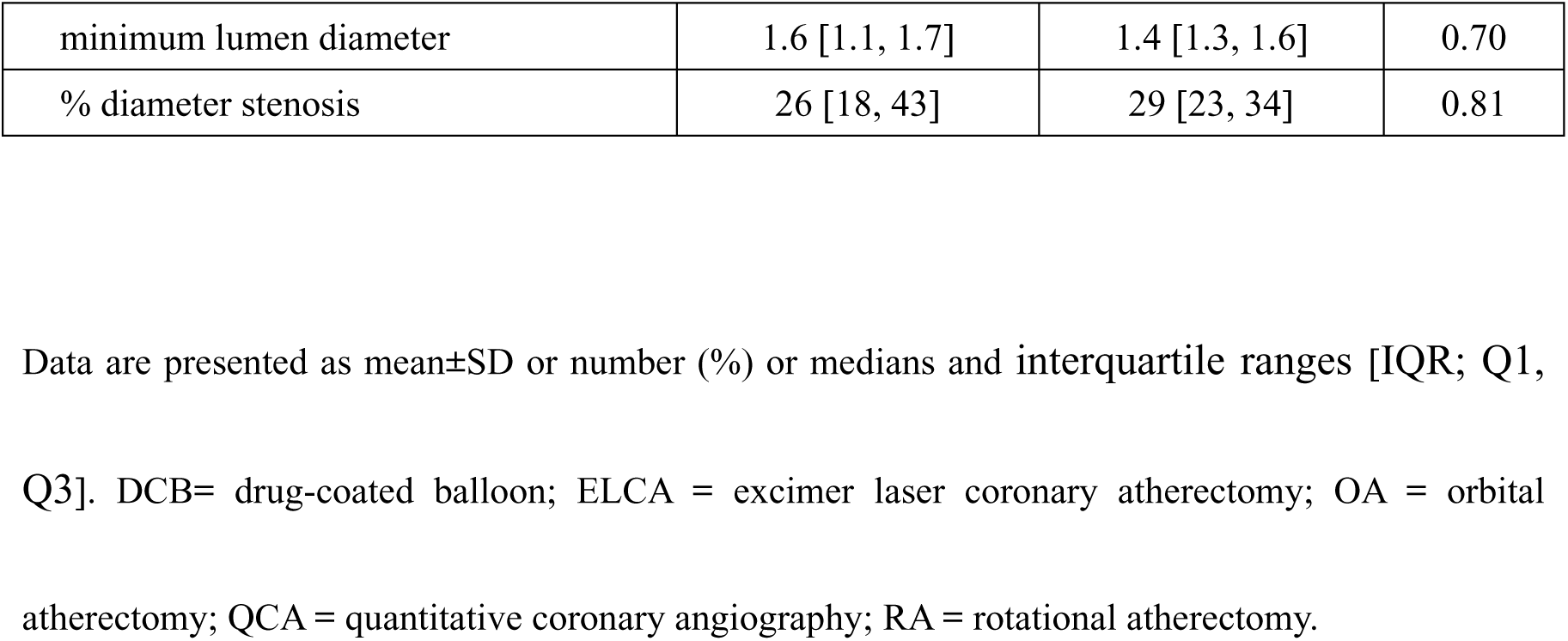
Procedural characteristics and QCA data.

### NIRS-IVUS FINDINGS

The timing of follow-up catheterization did not differ significantly between the two groups (250 days [IQR; 231, 252] in the lipid-rich group vs. 245 days [IQR; 231, 261] in the non-lipid-rich group; P = 0.66). Serial NIRS-IVUS data are presented in Table 3. Mean lumen volume was smaller in the lipid-rich group than in the non-lipid-rich group immediately after the procedure, and this difference remained consistent at the 8-month follow-up. Regarding the serial changes in each volume, the incidence of LLE did not differ significantly between the two groups (27 % in the lipid-rich group vs. 39 % in the non-lipid-rich group; P = 0.52). In contrast, percent plaque volume showed minimal change in the lipid-rich group (+0.3%), while a significant reduction was observed in the non-lipid-rich group (−3.6%; P = 0.045). Consequently, percent plaque volume at the 8-month follow-up was significantly higher in the lipid-rich group (54 % vs. 47 %; P = 0.015). Additionally, mean vessel volume tended to increase in the lipid-rich group compared with the non-lipid-rich group (+0.34 mm^3^/mm vs. −0.07 mm^3^/mm; P = 0.11). Notably, focusing on plaque composition, mean lipid plaque volume significantly decreased in the lipid-rich group compared with the non-lipid-rich group (−0.68 mm^3^/mm vs. −0.16 mm^3^/mm; P = 0.0020), whereas mean non-lipid plaque volume significantly increased (+0.87 mm^3^/mm vs. +0.08 mm^3^/mm; P = 0.020). maxLCBI_4_mm significantly decreased in the lipid-rich group compared with the non-lipid-rich group (−240 vs. −42; P = 0.0030), and the values at the 8-month follow-up were comparable between the two groups (224 vs. 152; P = 0.13).

**Table 3.**
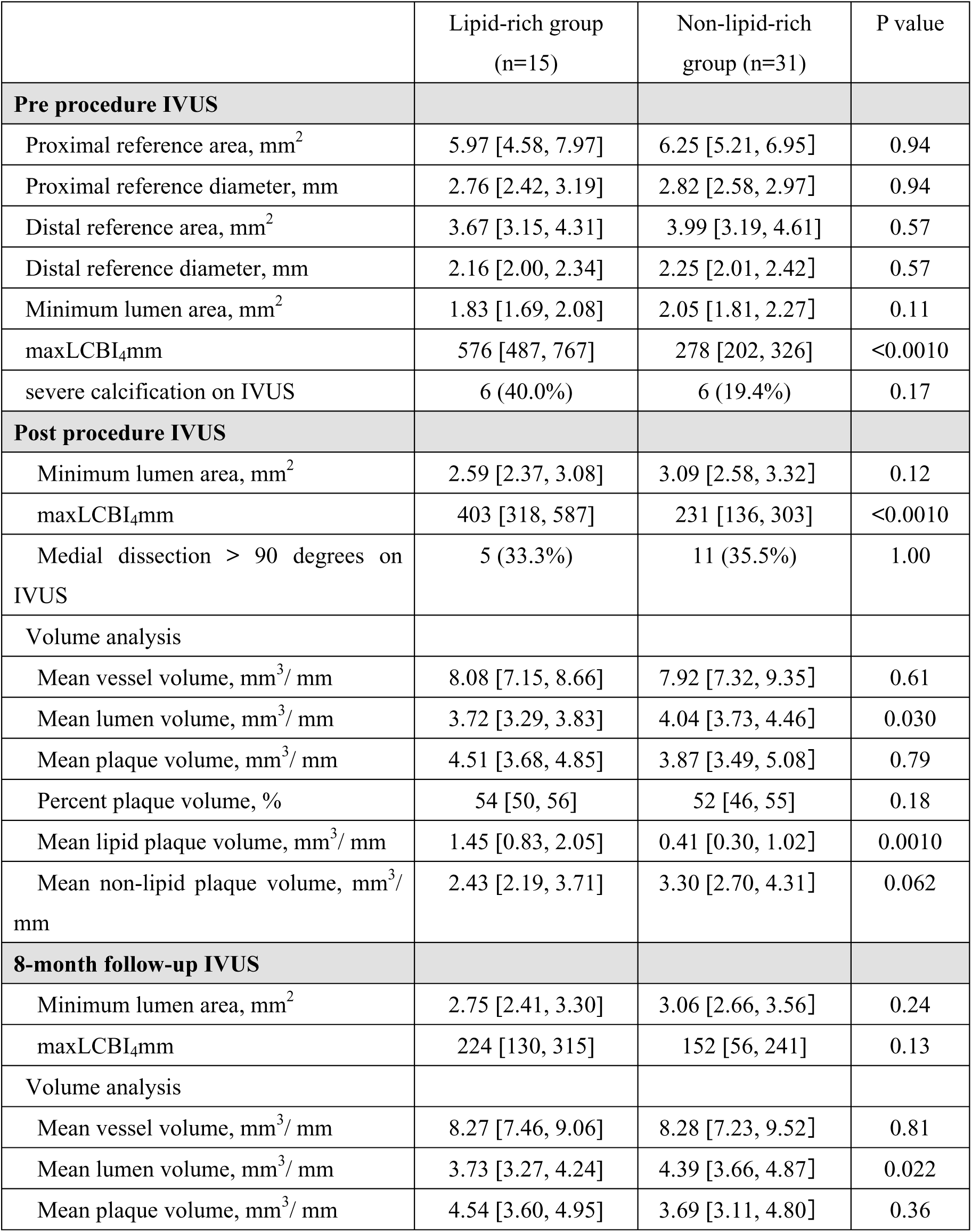
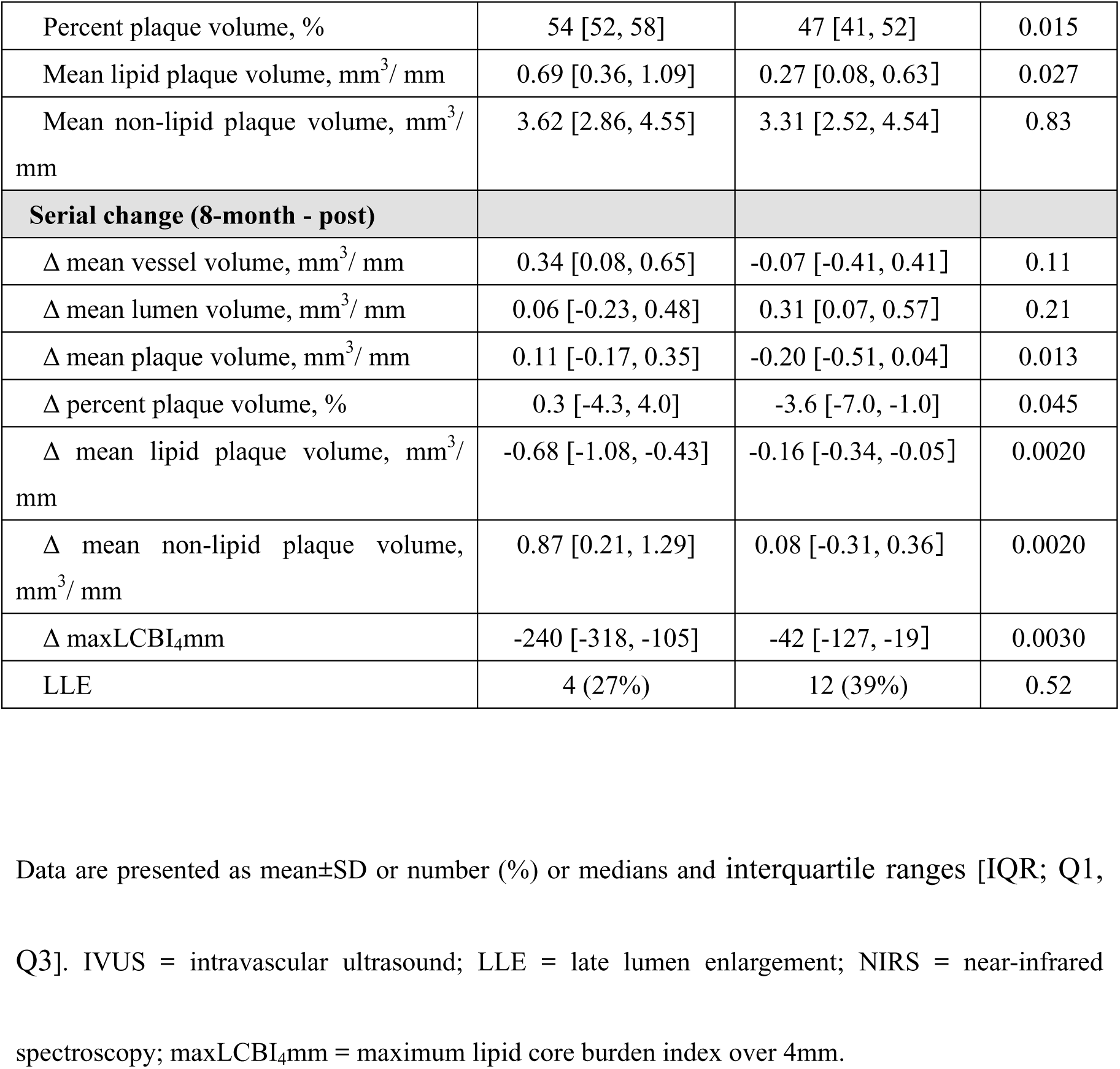
Serial NIRS-IVUS data.

### PREDICTORS OF LLE

Table 4 shows the results of univariate and multivariate analyses of predictors of LLE. High inflation pressure of the DCB and the presence of medial dissection > 90 degrees on IVUS immediately after the procedure were identified as significant predictors of LLE.

**Table 4.**
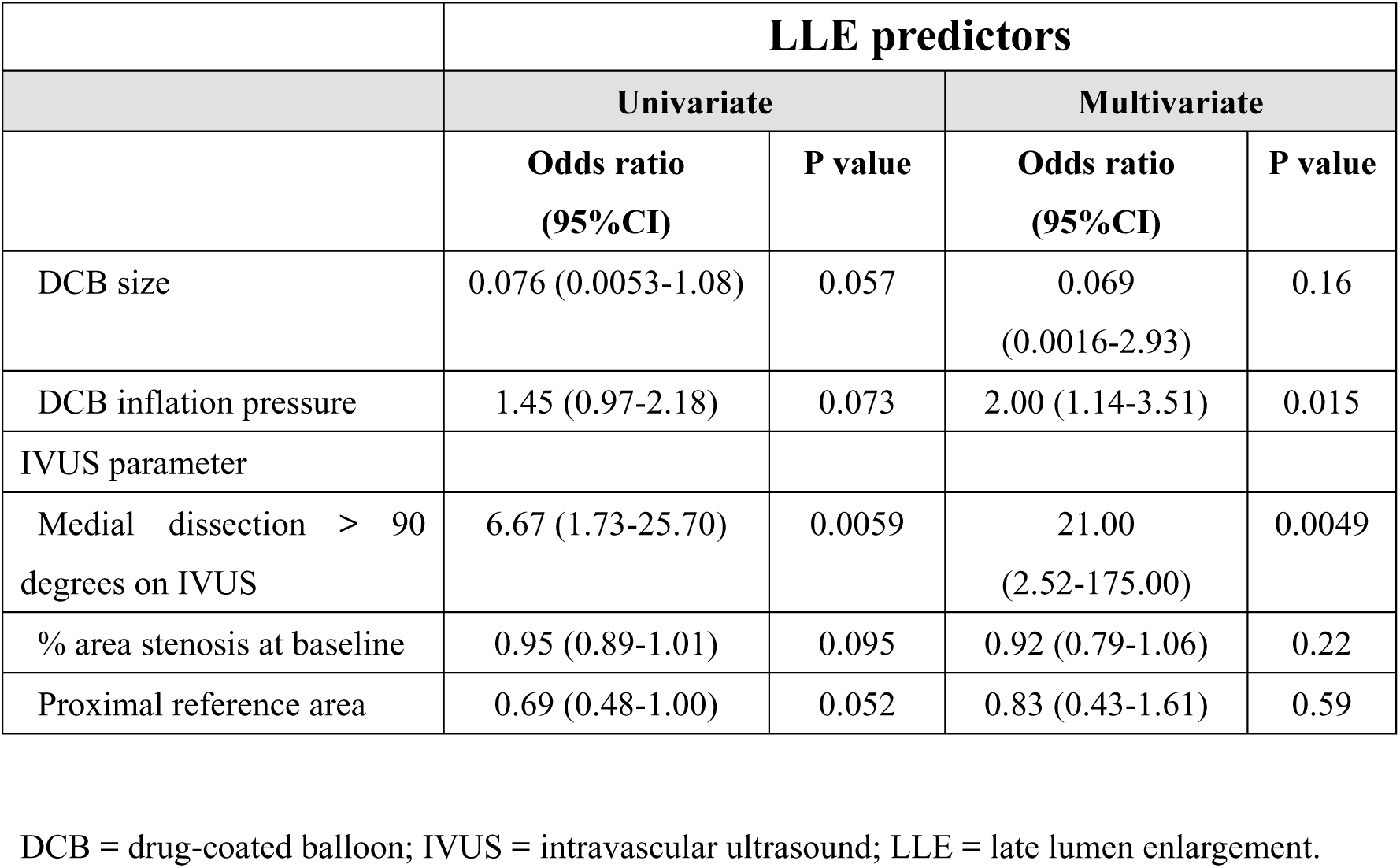
Univariate and multivariate analysis of predictors of LLE.

|  | **LLE predictors** |  |  |  |
| --- | --- | --- | --- | --- |
|  | **Univariate** |  | **Multivariate** |  |
|  | **Odds ratio**<br>**(95%CI)** | **P value** | **Odds ratio**<br>**(95%CI)** | **P value** |
| DCB size | 0.076 (0.0053-1.08) | 0.057 | 0.069<br>(0.0016-2.93) | 0.16 |
| DCB inflation pressure | 1.45 (0.97-2.18) | 0.073 | 2.00 (1.14-3.51) | 0.015 |
| IVUS parameter |  |  |  |  |
| Medial dissection > 90<br>degrees on IVUS | 6.67 (1.73-25.70) | 0.0059 | 21.00<br>(2.52-175.00) | 0.0049 |
| % area stenosis at baseline | 0.95 (0.89-1.01) | 0.095 | 0.92 (0.79-1.06) | 0.22 |
| Proximal reference area | 0.69 (0.48-1.00) | 0.052 | 0.83 (0.43-1.61) | 0.59 |

DCB = drug-coated balloon; IVUS = intravascular ultrasound; LLE = late lumen enlargement.

## Discussion

Our study revealed the following findings: 1) the incidence of LLE was comparable between the lipid-rich and non-lipid-rich groups following DCB angioplasty; 2) plaque regression was significantly less frequent in the lipid-rich group.; and 3) the lipid-rich groups demonstrated favorable plaque modification, including a reduction in lipid burden, resulting in comparable maxLCBI4mm values at the 8-month follow-up.

### VASCULAR RESPONSES FOLLOWING DCB ANGIOPLASTY

Paclitaxel coated on the DCB is an anti-inflammatory and antiproliferative drug.^16^ An animal study demonstrated that DCBs exert anti-inflammatory effects. These effects led to a histologically smaller accumulation of macrophages and smooth muscle cells, thereby halting plaque progression and, moreover, inducing plaque regression compared to conventional balloon therapy. The study concluded that DCB could serve as a regional endovascular therapy for atherosclerosis by reducing inflammation and slowing plaque progression.^16^ Moreover, another animal study revealed that DCB use was associated with decreased vessel wall fibrosis and a lower degree of constrictive vessel remodeling.^17^ Therefore, LLE following DCB angioplasty appears to result from two mechanisms: plaque regression and vessel enlargement. ^5^ It has also been reported that drug efficacy is enhanced by transmural distribution, and that extensive medial dissection may facilitate deeper drug penetration into the vessel wall.^4,18^ Indeed, medial dissection > 90 degrees was a strong predictor of LLE in the present study, consistent with previous findings.^4,14^

### LIPID-RICH LESIONS AND DCB ANGIOPLASTY

Notably, our study revealed that the incidence of LLE was comparable between the lipid-rich and non–lipid-rich groups. However, overall plaque regression was less frequently observed in the lipid-rich group. One possible explanation for this phenomenon is that mean vessel volume tended to increase in the lipid-rich group compared with the non–lipid-rich group. These findings suggest that the underlying mechanisms of LLE may differ between lipid-rich and non-lipid-rich lesions. Previous studies have demonstrated that plaque regression frequently follows plaque stabilization and that the degree of regression may be associated with plaque severity.^19,20,21,22^

Accordingly, plaque regression in our study occurred predominantly in the non– lipid-rich groups. In contrast, the reason why vessel enlargement tended to occur more frequently in the lipid-rich groups remains unclear. However, we speculate that in lipid-rich lesions, paclitaxel may penetrate more deeply into the vessel wall due to its high lipophilicity, thereby contributing primarily to vessel enlargement.^23^

It is also noteworthy that, in the lipid-rich group, the proportions of lipid and non-lipid plaques changed more dynamically compared with those in the non-lipid-rich group. Both mean lipid plaque volume and maxLCBI4mm showed significantly greater reductions in the lipid-rich group compared to the non-lipid-rich group, resulting in comparable maxLCBI_4_mm values between the two groups at the 8-month follow-up. Moreover, only two cases (13%) exhibited a maxLCBI_4_mm value over 400 at the 8-month follow-up, suggesting that plaque vulnerability was sufficiently stabilized in the lipid-rich groups.^7^ Although previous studies have shown that intensive lipid-lowering therapy contributes to the stabilization of lipid-rich plaques, we believe that the stabilization of plaque vulnerability observed in our study may be attributed to the effects of the DCB.^19,24^ Indeed, Veelen et al. reported the similar findings: maxLCBI_4_mm significantly decreased in the DCB treatment segments at the 9-month follow-up, while no such reduction was observed in non-DCB-treated segment.^25^ These results may indicate a direct effect of the DCB on the reduction of maxLCBI_4_mm.

### CLINICAL IMPLICATION

LLE is a specific and favorable vascular response in patients undergoing DCB angioplasty for de novo coronary artery disease. This study focused on baseline plaque characteristics, particularly the burden of lipid-rich plaques, and their association with LLE. The results revealed that LLE occurred to a similar extent in both lipid-rich and non-lipid-rich lesions. In contrast, the underlying mechanisms of LLE may differ between these two lesion types, with vessel enlargement predominantly associated with lipid-rich lesions. Currently both paclitaxel and sirolimus are used in DCBs, and comparable clinical outcomes have been reported.^23,26^ However, paclitaxel is highly lipophilic and has been reported to penetrate more deeply into the vessel wall compared with sirolimus.^27^ Therefore, these specific characteristics of paclitaxel can contribute to the favorable LLE outcomes observed in lipid-rich lesions.

Our data also demonstrated sufficient plaque stabilization in lipid-rich lesions at the 8-month follow-up. Lipid-rich plaques, widely recognized as vulnerable plaques, are strongly associated with adverse cardiac events.^7^ Accordingly, in addition to aggressive lipid-lowering therapy, stent implantation is occasionally performed even for non-culprit vulnerable plaques.^8,24,28^ Our findings suggest that DCB angioplasty may represent a promising therapeutic option for the management of vulnerable plaques, including those in non-culprit lesions. Compared with optimal medical therapy alone, DCB angioplasty may provide greater lumen gain and enhanced plaque stabilization without the need for a permanent metallic implant. Further large-scale studies are warranted to determine whether DCB angioplasty yields outcomes comparable to or superior to those achieved with optimal medical therapy or current drug-eluting stents in patients with non-culprit vulnerable plaques.

## Limitations

This study has several limitations. First, it is a pilot study with a relatively small sample size and no sample size calculation. In addition, the optimal cut-off value of maxLCBI_4_mm for identifying lipid-rich lesions in the context of DCB angioplasty remains unclear. Previous reports have proposed various cut-off values: maxLCBI_4_mm > 500 was useful for identifying the risk of peri-procedural myocardial infarction after stent implantation, and maxLCBI_4_mm ≥ 325 was also useful for identifying the future coronary events in non-target lesions.^6,29^ It has also been reported that maxLCBI_4_mm > 400 was useful for identifying vulnerable lipid-rich lesions, and this cut-off value was used in the present study.^7^ Nevertheless, our data suggest that the cut-off value (maxLCBI_4_mm > 400) used in this study may serve as a potential threshold for identifying lipid-rich lesions in the context of DCB angioplasty. Further investigation is needed to determine the optimal cut-off value of maxLCBI_4_mm in patients undergoing DCB angioplasty for de novo coronary artery disease.

### Conclusions

Despite limited plaque regression, lipid-rich lesions demonstrated favorable vascular responses, including LLE and plaque stabilization, following DCB angioplasty.

## Data Availability

Data are available from the corresponding author upon reasonable request.

## Acknowledgements

The authors thank all Osaka Rosai Hospital members who cooperated with the study.

## IRB information

This study was approved by the Medical Ethics Committee of Osaka Rosai Hospital (approval number: 2020-103) and the Ethics Committee of Osaka University (approval number: K25068).

---

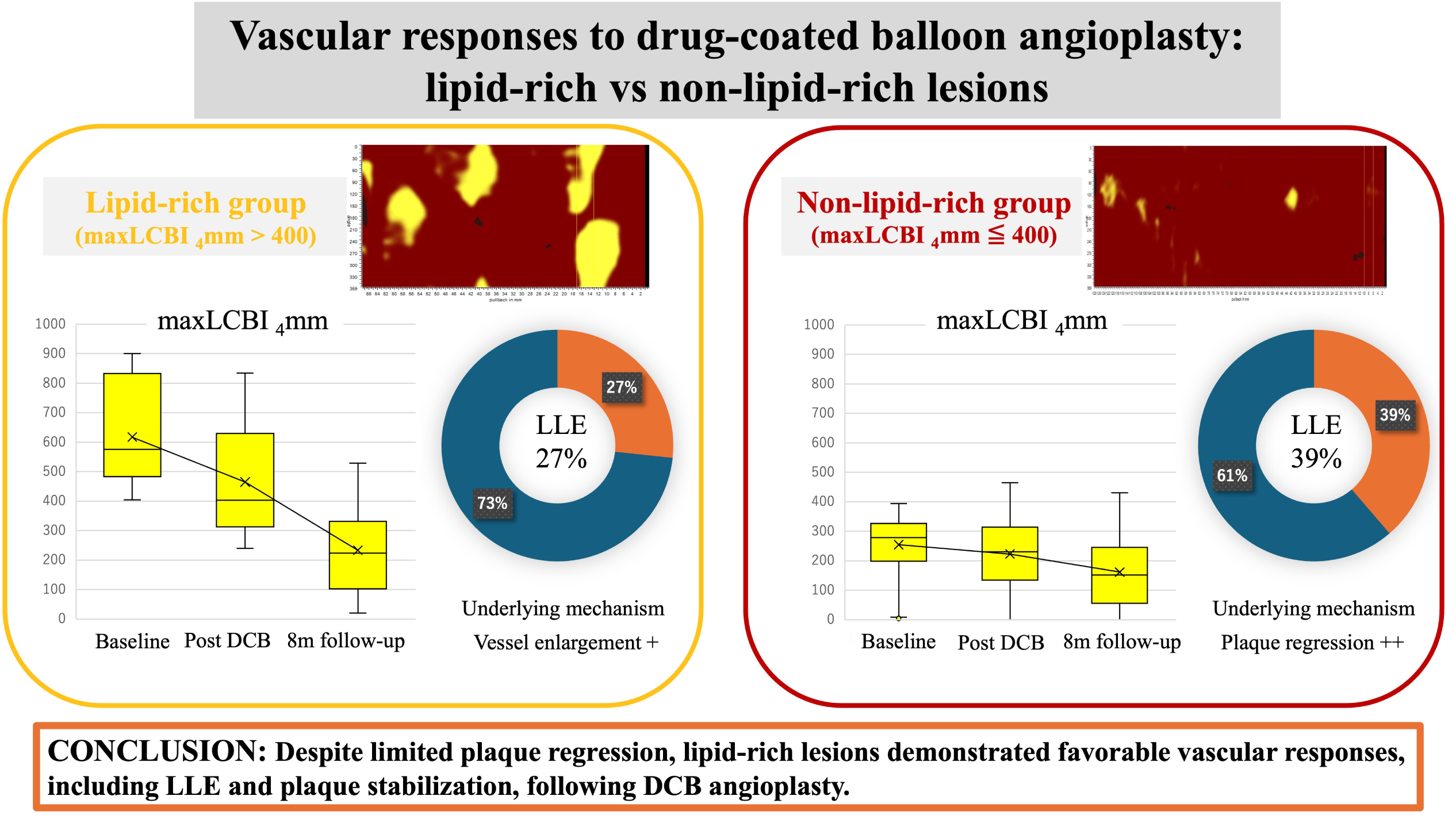

## ABBREVIATIONS LIST

DCB: drug-coated balloon
LLE: late lumen enlargement
maxLCBI4mm: maximum lipid core burden index over 4mm
NIRS-IVUS: near-infrared spectroscopy combined with intravascular ultrasound
QCA: quantitative coronary angiography

## References

1. Jeger RV, Farah A, Ohlow MA, Mangner N, Möbius-Winkler S, Leibundgut G, et al. Drug-coated balloons for small coronary artery disease (basket-small 2): An open-label randomised non-inferiority trial. Lancet. 2018;392:849–856

2. Gao C, He X, Ouyang F, Zhang Z, Shen G, Wu M, et al. Drug-coated balloon angioplasty with rescue stenting versus intended stenting for the treatment of patients with de novo coronary artery lesions (rec-cagefree i): An open-label, randomised, non-inferiority trial. Lancet. 2024;404:1040–1050

3. Yerasi C, Case BC, Forrestal BJ, Torguson R, Weintraub WS, Garcia-Garcia HM, et al. Drug-coated balloon for de novo coronary artery disease: Jacc state-of-the-art review. J Am Coll Cardiol. 2020;75:1061–1073

4. Yamamoto T, Sawada T, Uzu K, Takaya T, Kawai H, Yasaka Y. Possible mechanism of late lumen enlargement after treatment for de novo coronary lesions with drug-coated balloon. Int J Cardiol. 2020;321:30–37

5. Scheller B, Gemeinhardt O, Kleber FX. Late lumen enlargement after treatment of de-novo lesions with drug coated balloon catheters - glagov effect or plaque regression? Int J Cardiol. 2021;329:79–81

6. Erlinge D, Maehara A, Ben-Yehuda O, Bøtker HE, Maeng M, Kjøller-Hansen L, et al. Identification of vulnerable plaques and patients by intracoronary near-infrared spectroscopy and ultrasound (prospect ii): A prospective natural history study. Lancet. 2021;397:985–995

7. Waksman R, Di Mario C, Torguson R, Ali ZA, Singh V, Skinner WH, et al. Identification of patients and plaques vulnerable to future coronary events with near-infrared spectroscopy intravascular ultrasound imaging: A prospective, cohort study. Lancet. 2019;394:1629–1637

8. Park SJ, Ahn JM, Kang DY, Yun SC, Ahn YK, Kim WJ, et al. Preventive percutaneous coronary intervention versus optimal medical therapy alone for the treatment of vulnerable atherosclerotic coronary plaques (prevent): A multicentre, open-label, randomised controlled trial. Lancet. 2024;403:1753–1765

9. Muramatsu T, Kozuma K, Tanabe K, Morino Y, Ako J, Nakamura S, et al. Clinical expert consensus document on drug-coated balloon for coronary artery disease from the japanese association of cardiovascular intervention and therapeutics. Cardiovasc Interv Ther. 2023;38:166–176

10. Kleber FX, Rittger H, Bonaventura K, Zeymer U, Wöhrle J, Jeger R, et al. Drug-coated balloons for treatment of coronary artery disease: Updated recommendations from a consensus group. Clin Res Cardiol. 2013;102:785–797

11. Huber MS, Mooney JF, Madison J, Mooney MR. Use of a morphologic classification to predict clinical outcome after dissection from coronary angioplasty. Am J Cardiol. 1991;68:467–471

12. Jeger RV, Eccleshall S, Wan Ahmad WA, Ge J, Poerner TC, Shin ES, et al. Drug-coated balloons for coronary artery disease: Third report of the international dcb consensus group. JACC Cardiovasc Interv. 2020;13:1391–1402

13. Zhang M, Matsumura M, Usui E, Noguchi M, Fujimura T, Fall KN, et al. Intravascular ultrasound-derived calcium score to predict stent expansion in severely calcified lesions. Circ Cardiovasc Interv. 2021;14:e010296

14. Yamamoto M, Hara H, Kubota S, Hiroi Y. Predictors of late lumen enlargement after drug-coated balloon angioplasty for de novo coronary lesions. EuroIntervention. 2024;20:602–612

15. Mintz GS, Nissen SE, Anderson WD, Bailey SR, Erbel R, Fitzgerald PJ, et al. American college of cardiology clinical expert consensus document on standards for acquisition, measurement and reporting of intravascular ultrasound studies (ivus). A report of the american college of cardiology task force on clinical expert consensus documents. J Am Coll Cardiol. 2001;37:1478–1492

16. Chowdhury MM, Singh K, Albaghdadi MS, Khraishah H, Mauskapf A, Kessinger CW, et al. Paclitaxel drug-coated balloon angioplasty suppresses progression and inflammation of experimental atherosclerosis in rabbits. JACC Basic Transl Sci. 2020;5:685–695

17. Pavo N, Samaha E, Sabdyusheva I, von Strandmann RP, Stahnke S, Plass CA, et al. Coating of intravascular balloon with paclitaxel prevents constrictive remodeling of the dilated porcine femoral artery due to inhibition of intimal and media fibrosis. J Mater Sci Mater Med. 2016;27:131

18. Hou D, Rogers PI, Toleikis PM, Hunter W, March KL. Intrapericardial paclitaxel delivery inhibits neointimal proliferation and promotes arterial enlargement after porcine coronary overstretch. Circulation. 2000;102:1575–1581

19. Dohi T, Maehara A, Moreno PR, Baber U, Kovacic JC, Limaye AM, et al. The relationship among extent of lipid-rich plaque, lesion characteristics, and plaque progression/regression in patients with coronary artery disease: A serial near-infrared spectroscopy and intravascular ultrasound study. Eur Heart J Cardiovasc Imaging. 2015;16:81–87

20. Daoud AS, Jarmolych J, Augustyn JM, Fritz KE. Sequential morphologic studies of regression of advanced atherosclerosis. Arch Pathol Lab Med. 1981;105:233–239

21. Williams KJ, Feig JE, Fisher EA. Cellular and molecular mechanisms for rapid regression of atherosclerosis: From bench top to potentially achievable clinical goal. Curr Opin Lipidol. 2007;18:443–450

22. Hirayama A, Saito S, Ueda Y, Takayama T, Honye J, Komatsu S, et al. Qualitative and quantitative changes in coronary plaque associated with atorvastatin therapy. Circ J. 2009;73:718–725

23. Sedhom R, Hamed M, Elbadawi A, Mohsen A, Swamy P, Athar A, et al. Outcomes with limus-vs paclitaxel-coated balloons for percutaneous coronary intervention: Meta-analysis of randomized controlled trials. JACC Cardiovasc Interv. 2024;17:1533–1543

24. Räber L, Ueki Y, Otsuka T, Losdat S, Häner JD, Lonborg J, et al. Effect of alirocumab added to high-intensity statin therapy on coronary atherosclerosis in patients with acute myocardial infarction: The pacman-ami randomized clinical trial. Jama. 2022;327:1771–1781

25. van Veelen A, Küçük IT, Garcia-Garcia HM, Fuentes FH, Kahsay Y, Delewi R, et al. Paclitaxel-coated balloons for vulnerable lipid-rich plaques. EuroIntervention. 2024;20:e826–e830

26. Ahmad WAW, Nuruddin AA, Abdul Kader M, Ong TK, Liew HB, Ali RM, et al. Treatment of coronary de novo lesions by a sirolimus-or paclitaxel-coated balloon. JACC Cardiovasc Interv. 2022;15:770–779

27. Aihara K, Torii S, Ito M, Koseki K, Shiozaki M, Sato Y, et al. Biological differences of three paclitaxel- and sirolimus-coated balloons on coronary lesions in a rabbit model. EuroIntervention. 2024;20:e389–e398

28. Stone GW, Maehara A, Ali ZA, Held C, Matsumura M, Kjøller-Hansen L, et al. Percutaneous coronary intervention for vulnerable coronary atherosclerotic plaque. J Am Coll Cardiol. 2020;76:2289–2301

29. Goldstein JA, Maini B, Dixon SR, Brilakis ES, Grines CL, Rizik DG, et al. Detection of lipid-core plaques by intracoronary near-infrared spectroscopy identifies high risk of periprocedural myocardial infarction. Circ Cardiovasc Interv. 2011;4:429–437

